# Transitions in cigarette and ENDS use in the PATH Study: a multistate transition model analysis of adults in 2021–2022 compared to previous years

**DOI:** 10.64898/2025.12.15.25342290

**Authors:** Andrew F. Brouwer, Olivia K. Roberts, Jihyoun Jeon, Evelyn Jimenez-Mendoza, Stephanie R. Land, Neal D. Freedman, Rossana Torres-Alvarez, Ritesh Mistry, David T. Levy, Rafael Meza

## Abstract

**Introduction:** Electronic nicotine delivery systems (ENDS) products continue to evolve, and so ongoing analysis of transition rates over time is important for tracking real-world associations between ENDS and cigarette use and for providing the information necessary to project future public health outcomes.

**Methods:** Using the Population Assessment of Tobacco and Health (PATH) Study Waves 6–7 (2021–2022), we applied a Markov multistate transition model to estimate transition rates for initiation and cessation of each product. We estimated one-year transition probabilities for each transition. These results were compared to estimated rates and probabilities in Waves 1–6 (2014– 2021).

**Results:** The fraction adopting ENDS use in 2021–22 among those who had never previously established tobacco product use, those not currently using tobacco products, and those currently smoking cigarettes increased to 0.5% (95% confidence intervals [CI]: 0.4, 0.6%), 2.7% (95% CI: 2.3, 3.2%), and 6.8% (95% CI: 6.1, 7.6%), respectively. These increases were driven by young adults (ages 18–24), with respective transition fractions of 2.7% (95% CI: 2.3, 3.2%), 23.6% (95% CI: 20.2, 27.0%), and 19.2% (95% CI: 14.0, 24.5%). The fraction of adults who transitioned from dual cigarette and ENDS use to cigarette-only use remained around 25% (26.2% [95% CI: 21.7, 30.7%]), while the fraction who transitioned to ENDS-only use increased to 24.2% (95%CI: 20.5, 27.9%). The increase in the dual to ENDS-only use transition was also driven by young adults (34.4% [95% CI: 26.2, 42.6%]) and adults ages 25–34 (29.4% [95% CI: 23.1, 35.7%]).

**Conclusion:** Public health efforts are needed to promote cigarette cessation among older adults, specifically.

**What this paper adds:** *What is already known on this topic:*

- Transitions in cigarette and ENDS use have been changing over time. Young adults have been early adopters of ENDS, with older adults less likely to try ENDS or to completely switch from cigarettes to ENDS.
- Frequency of product use likely impacts the likelihood of product quitting or switching.

*What this study adds:*

- We found increasing adoption of ENDS among adults who have never smoked, those not currently using cigarettes or ENDS, and those using cigarettes only. These patterns were driven by young adults, with little cigarette cessation or switching to ENDS among older adults.
- Daily (vs non-daily) use of ENDS facilitated cigarette cessation among those using cigarettes and ENDS, but it was a barrier to ENDS cessation among those using ENDS only.

*How this study might affect research, practice, or policy:*

- Public health efforts are needed to promote cigarette cessation among older adults who smoke, many of whom may already be experiencing the health effects of tobacco use.
- Studies are needed to develop strategies for leveraging ENDS to maximize smoking cessation while also helping those who successfully quit smoking to avoid long-term ENDS use.

## Introduction

The tobacco and nicotine product marketplace continues to evolve rapidly in the US. The US Food and Drug Administration authorized the marketing of the first electronic nicotine delivery systems (ENDS) in the US in October 2021. Meanwhile, the sale of oral nicotine pouches rose over the 2019–22 period,^1^ potentially competing with existing products or initiating new people to nicotine addiction who may begin using ENDS or cigarettes. These and other changes may have continued to shift patterns of use and transitions between ENDS and cigarettes. Ongoing surveillance of these transitions is important for understanding how the public health impact of ENDS may change over time and for updating projections of future impact on health effects.

We previously leveraged the Population Assessment of Tobacco and Health (PATH) Study, a nationally representative longitudinal survey of the US, using a multistate transition model to analyze how transitions among ENDS and cigarette use changed over Waves 1–6 (2014–21).^2–4^ In the present analysis, we extend our models to compare previous transitions to those observed between Waves 6 and 7 of PATH (2021–22). Our multistage transition model approach, which estimates the transition rates that underlie observed transitions between products,^5–14^ allows us to see how the rates of competing transitions result in observed transition probabilities. Directly estimating these underlying transition rates provides a clearer picture of important shifts in use patterns and produces parameters for tobacco use status quo and scenario simulation modeling that can be used to investigate the potential impact of regulations.

## Methods

### Data

The Population Assessment of Tobacco and Health (PATH) Study is a nationally representative longitudinal cohort study of tobacco and nicotine product use behaviors in the US among the civilian, non-institutionalized population. Our analysis compared 23,803 adults in the Wave 4 cohort in Wave 6 (March 2021 to November 2021, abbreviated as 2021) and Wave 7 (January 2022 to April 2023, abbreviated as 2022). Transitions between these waves were compared to transitions between each pair of previous waves, with particular attention to comparison to the transitions in the preceding period, from Wave 5 (December 2018 to November 2019, abbreviated 2019) to Wave 6. Characteristics of the population for each wave pair are given in Table S1. This analysis was not regulated as human subjects research (University of Michigan Institutional Review Board HUM00162265).

Follow-up time for participants was approximately one year between Waves 6 and 7 and was treated as exactly one year in the model. Follow-up between other pairs of Waves were assumed to be either one or two years, as appropriate for the corresponding wave pair. We categorized each participant’s ages as 18–24, 25–34, 35–54, or 55–90 years. Each participant’s product use was categorized as never established use (of both products), non-current use (of both products), cigarette-only use, ENDS-only use, or dual use of cigarettes and ENDS, as in previous work.^2–4^ Established cigarette use was defined as having smoked at least 100 cigarettes in one’s lifetime, and established ENDS use was defined as ever having “fairly regularly” used ENDS. Current use of cigarettes and ENDS was defined as ever established and any past 30-day use of that product, and current dual use was defined as established and current use of both cigarettes and ENDS. Non-current use was defined as no past-30-day use of either product by a participant who had previously established use of one or both products; because “former” use is generally understood to refer to longer-term cessation, we refer to “non-current” use to describe no past-30-day use. Under our definitions, current experimental (i.e., current but not established use) product use is not included in the corresponding current use group; for example, a participant who had smoked 100 cigarettes but who had never “fairly regularly” used ENDS and used both products in the past 30-days would be included in the “cigarette-only use” category.

### Transition modeling

We applied our multistate transition model to analyze the underlying transition hazard rates between the product use categories for adults overall and in each of the four age categories. Multistate transition models are finite-state, Markov stochastic process models that assume that transition hazard rates depend only on the current state and not on past states or transition history. Technical details of the multistate transition model are provided in the Supplemental Material. Using discrete-time observations, the model estimated the instantaneous risk of transition from one state to another, i.e., transition hazard rates, which collectively define the probability of transitioning from one state to any other at a future time.

For Waves 6–7, we incorporated Wave 4 longitudinal survey weights into the model, with replicate weights used to calculate variance and confidence intervals, as described in the PATH user guide.

However, preliminary model analysis using the default weights evidenced poor fits to the empirical transitions between ENDS and dual use, as discussed further in the Results. To address this problem, we renormalized the weights within product use categories, giving equal weight to each product use state in the model. This reweighting approach resulted in probabilities that better reflected the observed empirical transition fractions, as discussed in the Results. For consistency across the analyses, we re-estimated transitions for all previous waves with the renormalized weights, although re-normalization had little effect on most transitions in previous waves.

Using the estimated transition rates, we calculated the 1-year transition probabilities overall and for each age group in each time period separately. Transition probabilities account for the possibility of multiple transitions between product use states over the period. For Waves with a 2-year gap, the 1-year transition probabilities reflect estimated transition fractions per year over the 2-year period.

Additionally, we estimated associations between transition rates and non-daily vs daily use of each product, accounting for gender, age group, race and ethnicity, for each wave pair using the renormalized weights. All multistate transition models were estimated using the wmsm function^2^ in R (v1.3, publicly available at https://tcors.umich.edu/Resources_Research.php), which is an extension of the msm function^15^ modified to incorporate survey weights.

## Results

In Wave 6 (2021), the average weighted prevalence of never established use was 57.3%, the prevalence of non-current use was 25.3%, the prevalence of current cigarette-only use was 12.7%, the prevalence of current ENDS-only use was 3.2%, and the prevalence of current dual cigarette and ENDS use was 1.5%.

Weighted empirical transitions probabilities for Wave 6–7 (Fig S1A) were broadly captured by the model with the original weighting approach (Fig S1B), with the exception of the probability of transitions from dual use to ENDS-only use (empirical: 23.3%; estimated 28.4%) or remaining in dual use (empirical: 45.3%; estimated 39.2%), both of which were off by about 5 percentage points. The probabilities estimated using the new weighting approach with weights renormalized within product use categories (Fig S1C) better reflected the two transitions (24.4% and 43.9%, respectively). Re-estimation of transitions in previous wave pairs using the new weighting approach had little effect on transition estimates (Fig S2). All subsequent results report models using the new weighting approach.

Between 2019–21 and 2021–22, all transition rates statistically significantly increased, except for cigarette initiation, which significantly decreased, and the dual to cigarette-only use transition, which was not significantly different (Fig 1A). We plot one-year transition heatmaps for 2019–21 and 2021–22 in Fig 1B and C, and we visualize longer-term trends in transitions overall in Fig 2 and by age group in Fig 3. Cigarette initiation from never established use continued to decline from 0.2% to 0.1% (Fig 1C; Fig 2A), but ENDS initiation continued to increase to 0.5% (95% confidence intervals [CI]: 0.4, 0.6%) in 2021–22, up from 0.3% (95% CI: 0.3, 0.4%) in 2019–22. This change was driven by young adults, 2.7% (95% CI: 2.2, 3.2%) of whom initiated ENDS-only use in 2021–22 (Figure 3B). Small overall increases in other transitions representing adoption of ENDS use—from non-current to ENDS-only use (from 1.0% [95% CI: 0.8, 1.2%] in 2019–21 to 2.7% [95% CI: 2.3, 3.2%] in 2021–22; Fig 2B) and from cigarette-only to dual use (from 4.0% [95% CI: 3.6, 4.5%] to 6.8% [95% CI: 6.1, 7.6%]; Fig 2C)—were also driven by young adults. Among young adults, 23.6% (95% CI: 20.2, 27.0%) of those in the non-current use category transitioned to ENDS-only use in 2021–22 (Fig 3D), and 19.2% (14.0, 24.5%) of those using cigarettes only transitioned to dual use (Fig 3F), up from 9.2% (95% CI: 6.8, 11.7%) and 14.2% (95% CI: 11.5, 16.9%), respectively, in 2019–21.

**Figure 1:**
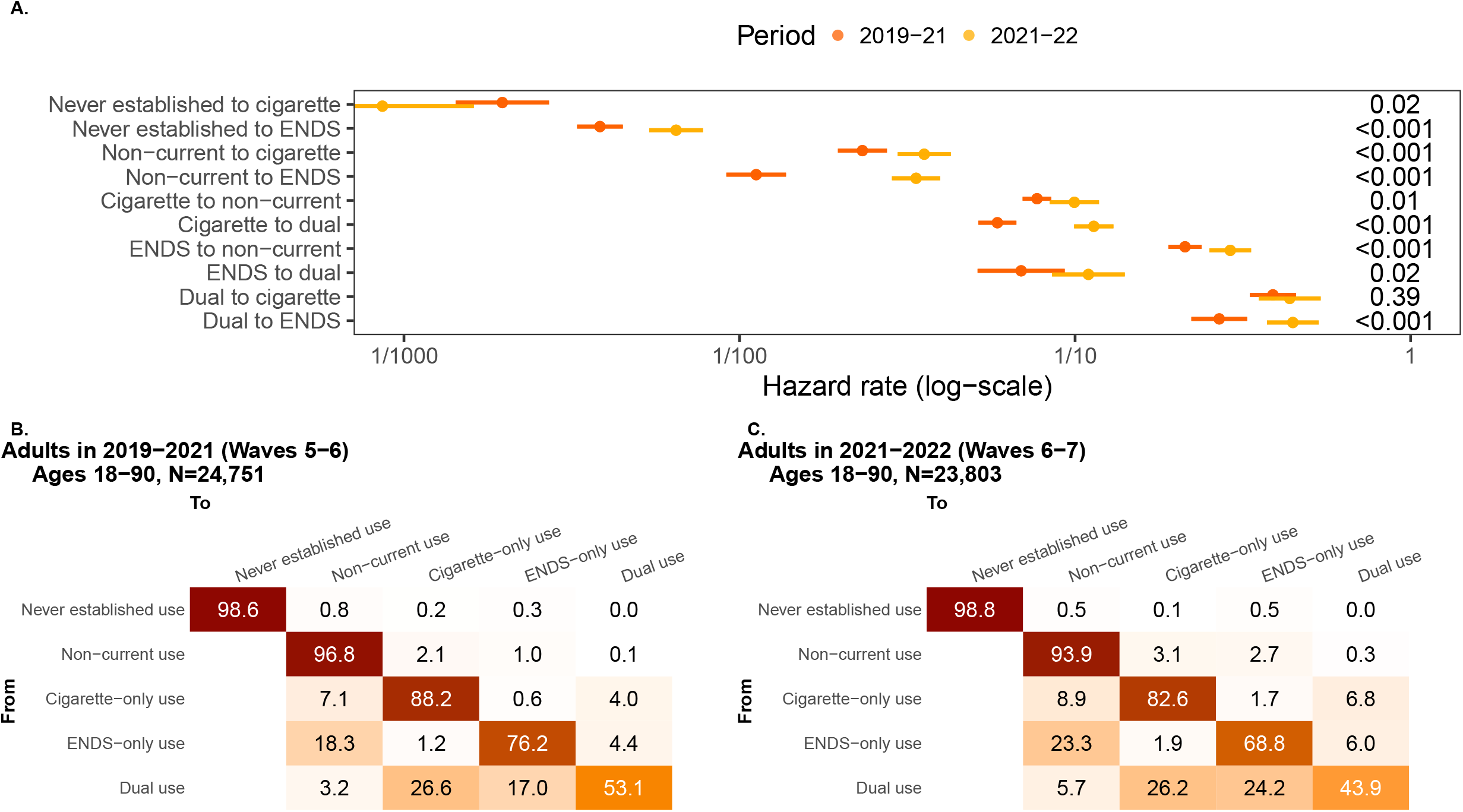
A) Transition hazard rates among adults in 2019–21 (Waves 5–6) and 2021–22 (Waves 6–7). A version of this figure including previous results from 2014–19 (Waves 1–5) is given in Fig S2. B) One-year transition probabilities for adults in 2019–21 (Waves 5-6), and C) 2021–22 (Waves 6-7). Confidence intervals for B and C are provided in Supplementary Table S2.

**Figure 2:**
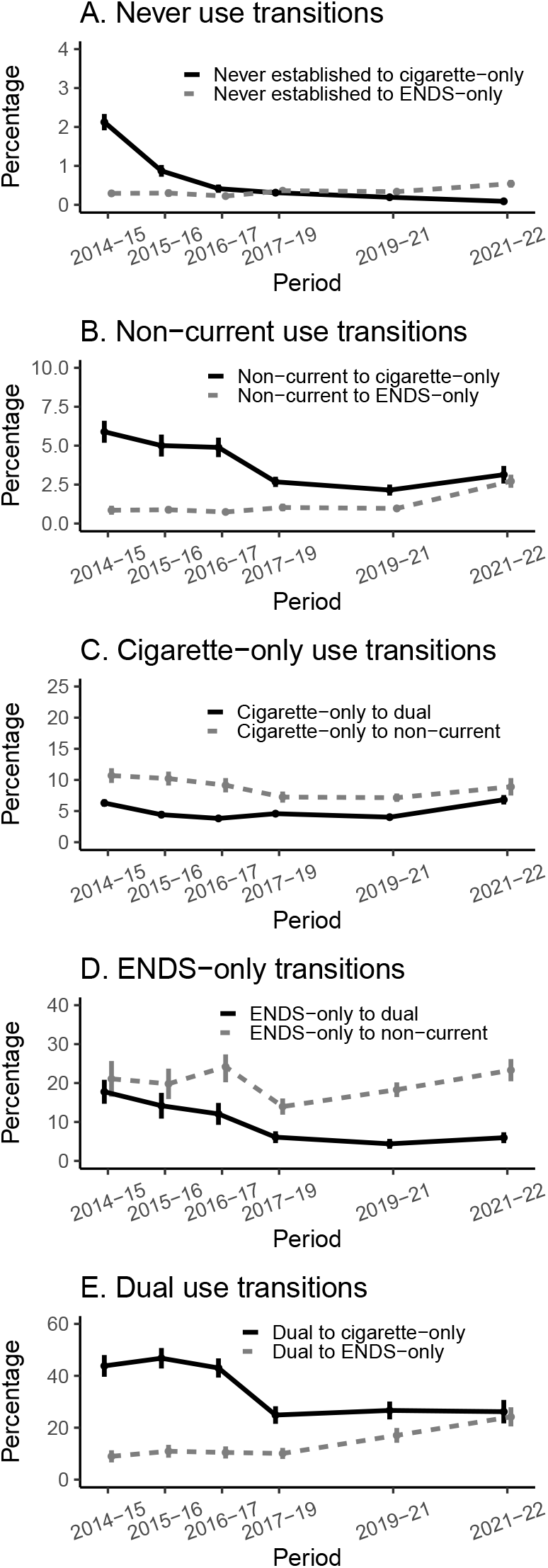
One-year transition probabilities and 95% confidence intervals for adults, PATH Waves 1-7 (2014–21), by product use state. Please note the change in y-axis range across product use categories. The x-axis is spaced by the year of the “from” Wave, and the y-axis is the modeled transition percentage between the pair of Waves. Point estimates and confidence intervals are provided in Supplementary Table S2.

**Figure 3:**
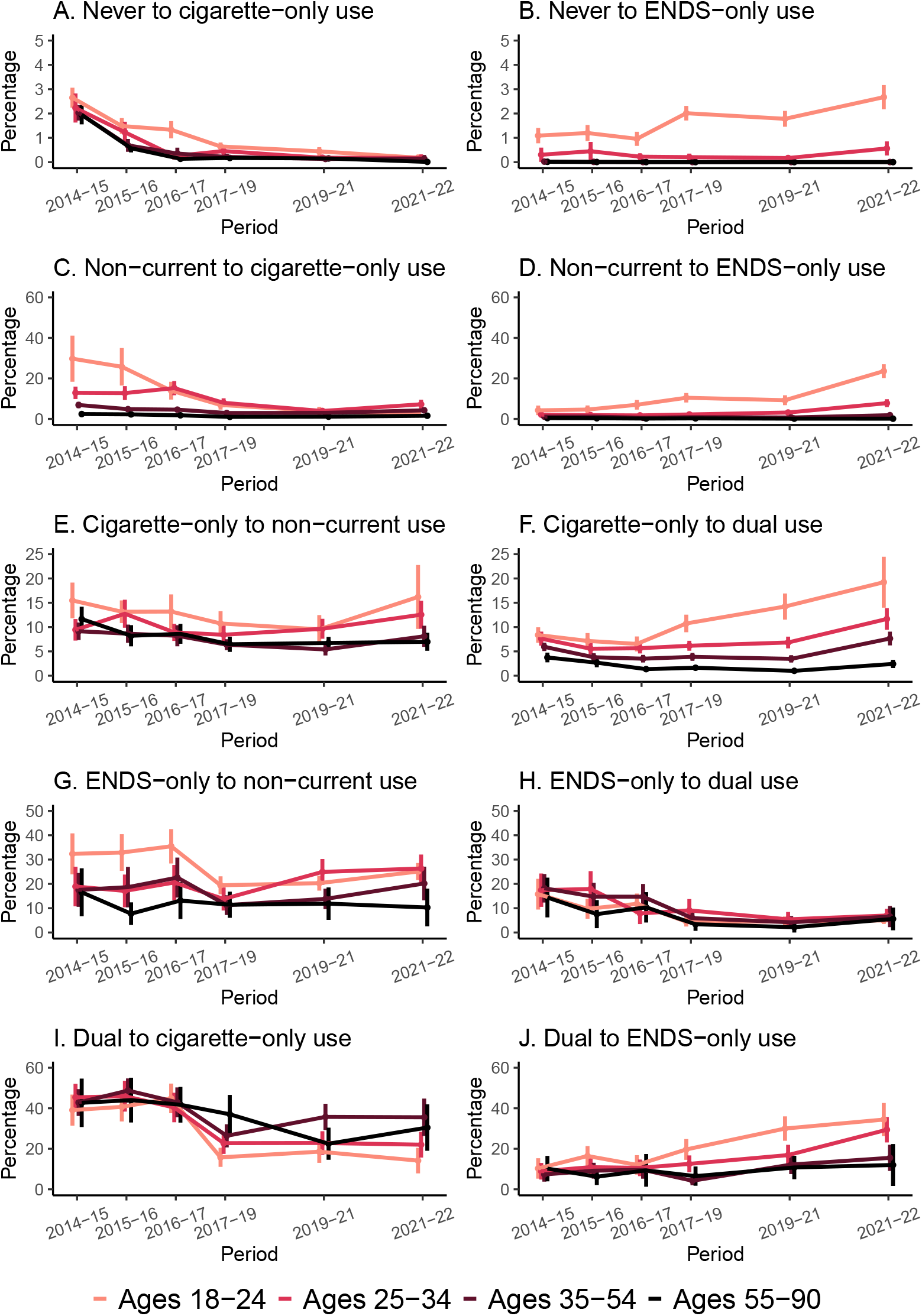
One-year transition probabilities and 95% confidence intervals for adults, PATH Waves 1-7 (2014–21), by transition and age group. Please note the change in y-axis range across product use categories. The x-axis is spaced by the year of the “from” Wave, and the y-axis is the modeled transition percentage between the pair of Waves. Point estimates and confidence intervals are provided in Supplementary Table S2.

Transitions from ENDS-only to dual use remained relatively low in 2021–22 (6.0%, 95% CI: 4.6, 7.4%; Fig 2D), with little difference by age group (Fig 3H). Transitions from ENDS-only to non-current use continued to increase, from 18.3% (95% CI: 16.4, 20.2%) in 2019–21 to 23.3% (95%CI: 20.5, 26.2%) in 2021–22 (Fig 2D). This transition probability was notably lower for older adults ages 55–90, at 11.9% (95% CI: 5.2, 18.5%) (Fig 3G).

Transitions from dual use to cigarette-only use remained approximately one quarter (26.6% [95% CI: 23.2, 30.1%) in 2019–21 and 26.2% [95% CI: 21.7, 30.7%] in 2021–22; Fig 2E). This transition was less common for young adults at 14.2% (95% CI: 7.9, 20.4%) and more common for older adults ages 55–90 (30.5% [95% CI: 19.0, 41.9%]; Fig 3I). The increase in transitions from dual to ENDS-only use (from 17.0% [95% CI: 14.2, 19.9%] in 2019–21 to 24.2% [95% CI: 20.5, 27.9%] in 2021–22), was driven by young adults (34.4% [95% CI: 26.2, 42.6%]; Fig 3J) and adults ages 25–34 (29.4% [95% CI: 23.1, 35.7%]).

Full results of the multivariable transitions models with transitions hazard ratios for product use frequency and the other covariates are given in the Supplementary Material (Table S2). For brevity, here we only highlight select results for product use frequency in 2021–22. Among adults who smoked cigarettes only, participants who smoked cigarettes non-daily had a higher rate of transitioning to non-current use (hazard ratio [HR] 6.54 [95% CI: 4.70, 9.09]) than those who smoked daily. Similarly, among those who used both ENDS and cigarettes, those who smoked cigarettes non-daily had a higher rate of transitioning to ENDS-only use (HR 3.07 [95% CI: 2.06, 4.57]) than those who smoked daily. The HRs by cigarette use frequency for the cigarette-only to dual use and from dual to cigarette-only use transitions were not statistically significant. Among adults who only vaped ENDS, those who vaped ENDS non-daily transitioned to non-current use at a higher rate (HR 2.63 [95% CI: 1.99, 3.45]) than who vaped daily, and, similarly, among those who used both products, those who vaped non-daily transitioned to ENDS-only use at a lower rate (HR: 0.59 [95% CI: 0.35, 0.98]) than those who vaped daily. The HRs by ENDS use frequency were not statistically significant for the ENDS-only to dual use or dual to cigarette-only use transitions.

## Discussion

In this analysis, we continued to track changes in transition rates between never, non-current, cigarette-only, ENDS-only, and dual use of both products among adults in the PATH Study, here adding transitions from Waves 6–7. In this period (2021–22), we observed increases in transitions representing the adoption of ENDS—from never established use (0.5%), from non-current use (2.7%), and from cigarette-only use (6.8% to dual use) compared to previous years. We observed marked differences by age group, with most increases in ENDS adoption driven by young adults (ages 18–24) and, to a lesser extent, adults ages 25–34. Similarly, the increase in transitions from dual to ENDS-only use (24.2%, catching up with transitions from dual to cigarette-only use, 26.2%), was again driven by the two younger age groups. Overall, there was comparatively little change for adults ages 35–54 and 55–90, where ENDS use and transitions to ENDS use remain low, and cigarette smoking cessation rates remained largely stagnant.

While ENDS are not harmless,^16–18^ they are likely less harmful than combustible cigarettes,^19–21^ and they have been promoted as cigarette cessation aids. Although we found an increasing propensity for younger adults to transition from cigarette-only to dual to ENDS-only use, both transitions along this pathway were comparatively rare for older adults. Specifically, in 2021–22, transitions from cigarette-only to dual use for older adults ages 55–90 were only 2.4% (95% CI: 1.6, 3.3%) compared to 19.2% (95% CI: 14.0, 24.5%) for younger adults, and transitions from dual use to ENDS-only for older adults were 12.0% (95% CI 1.7, 22.3%) compared to 34.4% (95% CI: 26.2, 42.6%) among younger adults. Because ENDS do not seem to be contributing to smoking cessation among older adults in a major way in the current marketplace, public health efforts are needed to promote cigarette cessation in older adults. Many studies have emphasized the need for health communication targeted to older adults specifically,^22–24^ as health communication strategies that work for young adults may not be successful for older adults.

Previous iterations of our transition surveillance have highlighted hazard ratios by sociodemographic factors^2^ and continuous age^3^ and how transition probabilities translate into numbers of people at the population level.^4^ In this analysis, we highlighted associations between non-daily vs daily use of cigarettes and ENDS. Unsurprisingly, we found that participants who used a product non-daily transitioned to non-current use at a greater rate than those who reported using that product daily, for both cigarettes (HR 6.54 [95% CI: 4.70, 9.09]) and ENDS (HR 2.62 [95% CI: 1.99, 3.45]). Among those who used both products and smoked cigarettes non-daily, transitions to ENDS-only use occurred at a greater rate (HR 3.07 [95% CI: 2.06, 4.57]) compared to those who smoked daily. Conversely, participants who vaped non-daily and smoked cigarettes transitioned to ENDS-only use at a lower rate (HR: 0.59 [95% CI: 0.35, 0.98]) than those who vaped daily. These results highlight an important challenge in promoting transitions from dual to ENDS-only to non-current use for people who smoke: successful transitions from dual to ENDS-only use are more likely among those frequently using ENDS, but frequent use is a barrier for the transition from ENDS-only to non-current use. Additional work is needed to better understand who might benefit from ENDS-mediated smoking cessation, especially if cessation through FDA-approved medication and counseling is not successful, and how to reduce the likelihood of long-term ENDS-only use after smoking cessation.

The strengths of this analysis include the use of the large, nationally representative longitudinal PATH Study data. Additionally, our use of multistate transition models allowed us to disentangle underlying transition rates from the observed transition fractions and explicitly account for non-uniform time between PATH Waves. However, our work did not account for participants’ use of other tobacco and nicotine products. Future work will need to consider how the introduction of oral nicotine pouches to the marketplace impacts transitions.

In this analysis, we found that transitions involving cigarettes and ENDS have continued to change over time for adults in the US. Many of the changes involving the adoption of ENDS or transitions from dual use of both products to ENDS-only use were driven by young adults, with little-to-no change among older adults. Public health efforts targeting older adults who smoke are needed to help this population to quit cigarettes, either by switching to ENDS-only or, ideally, completely quitting all nicotine products.

## Data Availability

The study only used publicly available data. Public Use Files from the Population Assessment of Tobacco and Health Study are available for download from an open access repository (https://doi.org/10.3886/ICPSR36498.v23).

## Acknowledgments

This project was funded through National Cancer Institute (NCI) and Food and Drug Administration (FDA) grant U54CA229974. The opinions expressed in this article are the authors’ own and do not reflect the views of the National Institutes of Health, the Department of Health and Human Services, or the United States government.

## Contributors

Conceptualisation and methodology: AFB and RMe; data curation: JJ and EJ-M; software: AFB, RTA, OKR; analysis and original draft preparation: AFB, OKR; review and editing: AFB, OKR, JJ, EJ-M, SRL, NF, RTA, RMi, DTL and RMe; funding acquisition: RMe and DTL; guarantor: AFB.

## Data availability statement

Public Use Files from the Population Assessment of Tobacco and Health Study are available for download from an open access repository (https://doi.org/10.3886/ICPSR36498.v23). Conditions of use are available on the aforementioned websites.

