## Supplementary material for "Transitions in cigarette and ENDS use in the PATH Study: a multistate transition model analysis of adults in 2021–2022 compared to previous years"

**Multistate transition modeling**

This technical appendix is reproduced in part from the supplementary material of Brouwer et al [6] but includes updated notation and technical details. Additional details may be found in Jackson [32], and a tutorial for working with the weighted multistate transition model (wmsm) code is available at <https://tcors.umich.edu/Resources_Research.php> along with the code itself.

A multistate transition model is a continuous-time, finite-state stochastic process with the Markov assumption that transition rates depend only on the current state and not on past states or transition history. We denote the state of the process at time $t$ as $S\left( t \right)$. We denote the probability that an individual is in state $j$ after an amount of $\Delta t$ since they were observed in state $i$ as

$$\begin{aligned} P_{ij}(t,t+\Delta t)=Pr\left[ S\left( t+\Delta t \right)=j | S\left( t \right)=i \right]. \#\left( 1 \right) \end{aligned}$$

In general, the transition probabilities can depend on the observation time $t$ in addition to the time span $\Delta t$, and in this analysis we consider discrete time periods over which we assume that that the model is homogeneous in time, i.e., $P_{ij}\left( t,t+\Delta t \right)= P_{ij}(0,\Delta t)$. Thus, we drop the dependence on $t$ moving forward and write $P_{ij}(\Delta t)$. We then define the hazard rate of the transition from state $i$ to state$j$, for $i\neq j$, as

$$\begin{aligned} q_{ij}=\lim_{\Delta t\to0} \frac{1}{\Delta t}Pr\left[ S\left( \Delta t \right)=j | S\left( 0 \right)=i \right]. \#\left( 2 \right) \end{aligned}$$

Transition hazards ratios $\rho_{ij,c_{l}}$may be determined for each transition $i$ to $j$ for each level $l$ of a covariate $c$, relative to the referent $c_{0}$: $q_{ij,c_{l}}=\rho_{ij,c_{l}}\times q_{ij,c_{0}}$. For time-varying covariates, a participant’s transition rate is determined by their characteristic at their most recent previous observation.

The transition hazard rates form a matrix $Q = \left[ q_{ij} \right],$ where the diagonal entries are given by $q_{ii}=-\sum_{j\neq i} q_{ij}$. The transition probability matrix $P\left( \Delta t \right)=\left[ P_{ij}\left( \Delta t \right) \right]$ is a function of the transition hazard rates and may be calculated as the matrix exponential of $\Delta t\cdot Q$, that is

$\begin{aligned} P\left( \Delta t \right)=\text{expm}\left( \Delta t\cdot Q \right).\#\left( 3 \right) \end{aligned}$

Transition probabilities can be estimate for any value of $\Delta t$, assuming that the transition hazards $q_{ij}$ do not change over that time period (assumption of homogeneity).

The values of the transition hazard rates $q_{ij}$of multistate transition model are estimated by maximizing a statistical likelihood $L$ given a set of observed states and times by comparing the observed states to the probabilities in $P\left( \Delta t \right)$ as a function of the transition hazard matrix $Q$. Specifically, consider a set of individuals $m = 1, \ldots,N$ and their observed states $s_{m,t_{m,k}}$at times $t_{m,k}$, where $k$ is the index of individual $m$’s $K_{m}$observations in the data. Denote the data as $s=\{s_{m,t_{m,k}}\}$. We assume the individuals are independent, and thus we multiply all the modeled probabilities of the observed transitions:

$$\begin{aligned} L\left( Q|s \right)=\prod_{m=1}^{N} \prod_{k=1}^{K_{m}-1} P_{s_{m,t_{m,k}},s_{m,t_{m,k+1}}}(t_{m,k+1}-t_{m,k}).\#\left( 4 \right) \end{aligned}$$

where $P_{i,j}$ is a function of $Q$ as in Eqn (3).

Participant weights $W_{m}$can be incorporated into a weighted likelihood $L^{*}$. Although it is not strictly necessary to normalize the weights to the population size, $w_{m}=N\cdot W_{m}/\sum_{v} W_{v}$, it is convenient to do so because the resulting likelihood will correspond to the unweighted likelihood when all participants have equal weight. The weighted likelihood is given by

$$\begin{aligned} L^{*}\left( Q|s \right)=\prod_{m=1}^{N} \prod_{k=1}^{K_{m}-1} \left( P_{s_{m,t_{m,k}},s_{m,t_{m,k+1}}}(t_{m,k+1}-t_{m,k}) \right)^{w_{m}}.\#\left( 5 \right) \end{aligned}$$

Following the msm package [32], we assume that estimated transition hazard rates $\hat{q}_{ij}$ are normally distributed on the log-scale, that is

$$\begin{aligned} \log\hat{q}_{ij}\sim N\left( \mu=\text{mean}\left( \log\hat{q}_{ij} \right),\sigma^{2}=V\left( \log\hat{q}_{ij} \right) \right),\#\left( 6 \right) \end{aligned}$$

where $V$ denotes the estimated variance.

We estimated weighted point estimates for the log transition hazard rates $\log\hat{q}_{ij}$ by minimizing ${-logL}^{*}\left( Q|s \right)$as a function of the transition hazard rates. (This approach is equivalent to maximum likelihood estimation).

Variance estimates $V\left( \log\hat{q}_{ij} \right)$are calculated using replicate weights $w_{m}^{r}$. Replicate weights are a way to account for complex survey design aspects, such as strata and primary sampling units. PATH uses a variant of balanced repeated replication called Fay’s method to calculate 100 replicate weights. We calculate $\log\hat{q}_{ij}^{r}$ for each $r$ as above. Then, we calculate the variance of $\log\hat{q}_{ij}^{r}$ as

$$\begin{aligned} V\left( \log\hat{q}_{ij} \right)=c\sum_{r=1}^{100} \left( \log\hat{q}_{ij}^{r}-\log\hat{q}_{ij} \right)^{2}\#\left( 7 \right) \end{aligned}$$

where $c=1/(100\left( 1-0.3 \right)^{2})$ as specified by PATH.

***Table S1****: Characteristics of adults in the Population Assessment of Tobacco and Health (PATH) study in 2014–15 (Waves 1–2), 2015–16 (Waves 2–3), 2016–17 (Waves 3–4), 2017–19 (Waves 4–5), 2019–21 (Waves 5–6), and 2021–22 (Waves 6–7) given as weighted percentages (%) and numbers (N). Note that in this table, “Tobacco & ENDS use state” is the prevalence for the earlier of the two waves.*

|  | Adults Waves 1–2 | | Adults Waves 2–3 | | Adults Waves 3–4 | | Adults Waves 4–5 | | Adults Waves 5–6 | | Adults Waves 6–7 | |
| --- | --- | --- | --- | --- | --- | --- | --- | --- | --- | --- | --- | --- |
|  | % | N | % | N | % | N | % | N | % | N | % | N |
| Total | 100 | 26,075 | 100 | 25,076 | 100 | 24,127 | 100 | 28,292 | 100 | 24,751 | 100 | 23,803 |
| Gender |  |  |  |  |  |  |  |  |  |  |  |  |
| Female | 52.2 | 13,218 | 52.2 | 12,880 | 52.1 | 12,538 | 52.0 | 14,676 | 52.0 | 13,075 | 52.1 | 12,656 |
| Male | 47.8 | 12,857 | 47.8 | 12,196 | 47.9 | 11,589 | 48.0 | 13,616 | 48.0 | 11,676 | 47.9 | 11,147 |
| Race/ethnicity |  |  |  |  |  |  |  |  |  |  |  |  |
| Non-Hispanic White | 64.9 | 15,606 | 64.7 | 14,780 | 64.4 | 13,950 | 63.7 | 15,932 | 63.2 | 13,733 | 62.9 | 12,772 |
| Non-Hispanic Black | 14.8 | 4,436 | 15.0 | 4,455 | 15.2 | 4,514 | 15.5 | 5,581 | 15.9 | 5,080 | 16.1 | 5.205 |
| Hispanic | 10.9 | 3,694 | 10.9 | 3,595 | 11.0 | 3,517 | 11.0 | 4,211 | 11.0 | 3,633 | 11.1 | 3,499 |
| Non-Hispanic Other/Unknown | 9.4 | 2,339 | 9.4 | 2,246 | 9.3 | 2,146 | 9.8 | 2,568 | 9.9 | 2,305 | 9.9 | 2,327 |
| Age (years) |  |  |  |  |  |  |  |  |  |  |  |  |
| 18-24 | 13.1 | 7,248 | 12.8 | 7,176 | 12.5 | 7,073 | 12.4 | 9,128 | 12.3 | 8,186 | 11.8 | 8,252 |
| 25-34 | 17.8 | 5,066 | 17.7 | 5,033 | 17.7 | 4,990 | 17.9 | 5,795 | 17.5 | 5,283 | 17.1 | 5,136 |
| 35-54 | 34.6 | 8,027 | 33.7 | 7,334 | 33.2 | 6,768 | 33.2 | 7,466 | 32.1 | 5,987 | 31.6 | 5,298 |
| 55-90 | 34.6 | 5,734 | 35.8 | 5,533 | 36.7 | 5,296 | 36.4 | 5,903 | 38.1 | 5,295 | 39.5 | 5,117 |
| Tobacco & ENDS use state |  |  |  |  |  |  |  |  |  |  |  |  |
| Never use | 61.2 | 12,570 | 58.4 | 11,689 | 57.1 | 11,379 | 58.0 | 14,138 | 57.7 | 13,023 | 57.3 | 12,897 |
| Non-current use | 20.1 | 4,001 | 22.1 | 4,442 | 23.4 | 4,582 | 23.0 | 5,308 | 23.7 | 4,829 | 25.3 | 5,182 |
| Cigarette-only use | 16.5 | 8,378 | 16.6 | 7,575 | 16.4 | 6,847 | 16.0 | 7,393 | 14.2 | 4,992 | 12.7 | 3,841 |
| Non-daily | 2.8 | 1,425 | 3.3 | 1,442 | 3.2 | 1,245 | 3.2 | 1,434 | 2.6 | 918 | 2.0 | 609 |
| Daily | 13.7 | 6,953 | 13.3 | 6,133 | 13.3 | 5,602 | 12.8 | 5,949 | 11.6 | 4,074 | 10.6 | 3,232 |
| ENDS-only use | 0.9 | 457 | 1.2 | 535 | 1.5 | 641 | 1.5 | 744 | 2.5 | 1,158 | 3.2 | 1,345 |
| Non-daily | 0.3 | 170 | 0.4 | 169 | 0.5 | 235 | 0.4 | 235 | 0.8 | 435 | 0.9 | 435 |
| Daily | 0.6 | 287 | 0.8 | 366 | 1.0 | 406 | 1.1 | 509 | 1.7 | 723 | 2.3 | 910 |
| Dual cigarette/ENDS user | 1.3 | 669 | 1.7 | 835 | 1.5 | 678 | 1.5 | 719 | 1.9 | 749 | 1.5 | 538 |
| Non-daily cigarette, non-daily ENDS | 0.1 | 67 | 0.2 | 97 | 0.2 | 88 | 0.2 | 98 | 0.3 | 111 | 0.2 | 78 |
| Non-daily cigarette, daily ENDS | 0.2 | 100 | 0.3 | 157 | 0.4 | 151 | 0.4 | 168 | 0.5 | 181 | 0.4 | 156 |
| Daily cigarette, non-daily ENDS | 0.6 | 332 | 0.8 | 406 | 0.7 | 293 | 0.6 | 316 | 0.7 | 291 | 0.6 | 196 |
| Daily cigarette, daily ENDS | 0.3 | 170 | 0.4 | 175 | 0.3 | 146 | 0.3 | 137 | 0.5 | 166 | 0.3 | 108 |

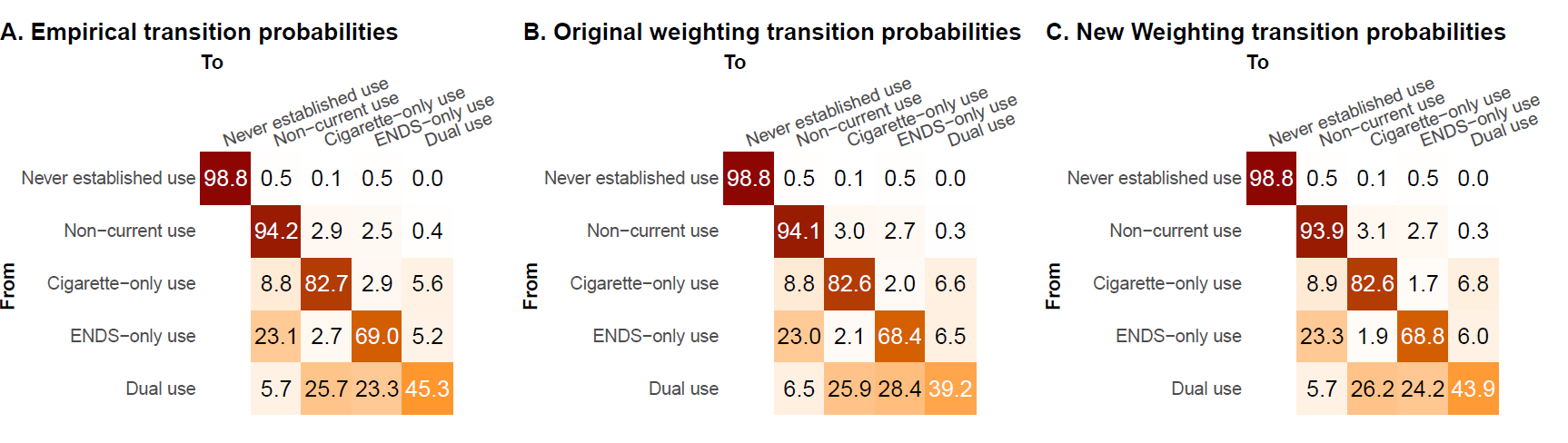

**Figure S1**: *A) Empirical transitions probabilities for PATH Waves 6–7 (2021–22). B) Modeled transition probabilities using the original Wave 4 cohort Wave 7 longitudinal weights. C) Modeled transition probabilities using the original Wave 4 cohort Wave 7 longitudinal weights, renormalized so that each product use state has equal weight.* *Our primary goal for the new weighting was to correct the transition probabilities from dual use, specifically the dual use to ENDS-only use transition and the dual use persistence fraction.*

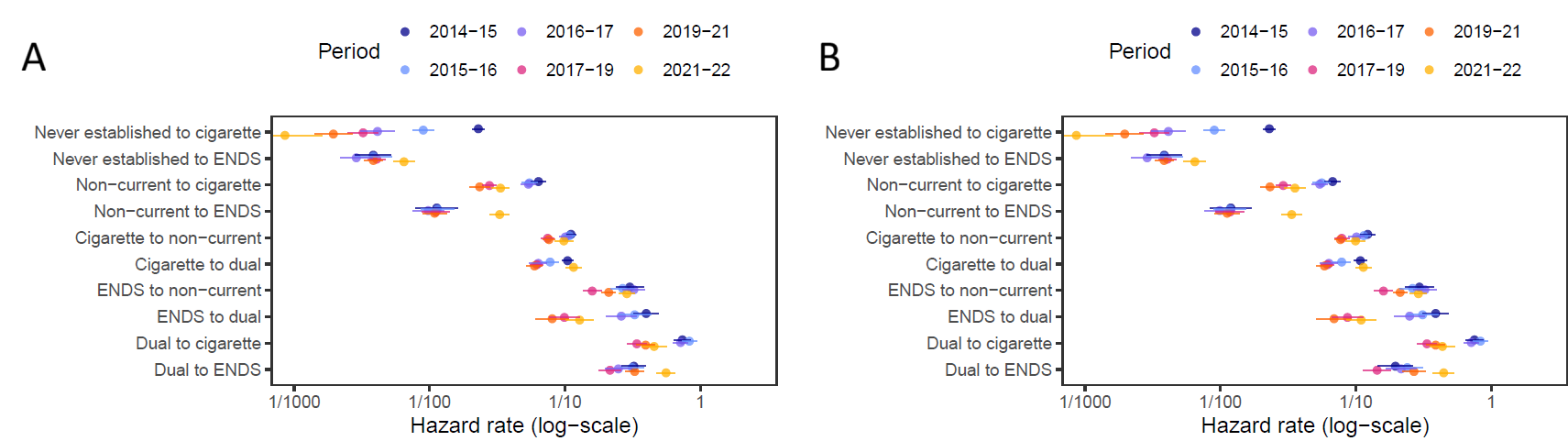

**Figure S2**: A) *Transition hazard rates using appropriate longitudinal among adults in each pair of PATH waves). B) Transition hazard rates re-estimated after renormalizing the weights to give equal weight to each product use state. The difference between the plots is primarily in the dual to ENDS use transition.*

**Table S2:** *One-year transition probabilities for each pair of PATH Waves overall and age group.*

Please find Supplementary Table S2.csv included separately.

**Table S3:** *Covariate hazard ratios for adults each pair of PATH Waves estimated by multivariable multistate transition models.*

|  | Male vs female | Age 18-24 vs 35-54 years | Age 25-34 vs 35-54 years | Age 55-90 vs 35-54 years | Non-Hispanic Black vs Non-Hispanic White | Hispanic vs Non-Hispanic White | Non-daily vs daily cigarette use | Non-Daily vs daily ENDS use |
| --- | --- | --- | --- | --- | --- | --- | --- | --- |
| Never to cigarette |  |  |  |  |  |  |  |  |
| Wave 1–2: 2014–15 | **1.84 (1.46, 2.31)** | 1.20 (0.85, 1.69) | 1.02 (0.69, 1.51) | 1.27 (0.92, 1.76) | **1.86 (1.17, 2.94)** | **2.39 (1.76, 3.24)** | — | — |
| Wave 2–3: 2015–16 | **1.99 (1.30, 3.05)** | **1.68 (1.01, 2.80)** | 1.66 (0.91, 3.03) | 0.90 (0.50, 1.60) | **3.36 (1.96, 5.76)** | **3.80 (2.14, 6.74)** | — | — |
| Wave 3–4: 2016–17 | 1.37 (0.75, 2.49) | **3.64 (1.55, 8.52)** | 0.57 (0.13, 2.51) | 0.44 (0.14, 1.34) | 2.19 (0.86, 5.53) | **2.58 (1.17, 5.67)** | — | — |
| Wave 4–5: 2017–19 | 1.25 (0.72, 2.16) | **2.96 (1.55, 5.63)** | 2.10 (0.83, 5.29) | 0.81 (0.35, 1.86) | 0.71 (0.35, 1.45) | 1.91 (0.98, 3.71) | — | — |
| Wave 5–6: 2019–21 | 2.09 (0.88, 4.96) | **3.26 (1.39, 7.69)** | 1.23 (0.32, 4.74) | 1.37 (0.49, 3.80) | 1.95 (0.49, 7.74) | **3.16 (1.20, 8.36)** | — | — |
| Wave 6–7: 2021–22 | 3.03 (0.65, 14.1) | 0.73 (0.07, 7.94) | 1.25 (0.19, 8.35) | 0.12 (0.01, 1.94) | **36.3 (2.12, 622.)** | **15.5 (2.12, 114.)** | — | — |
| Never to ENDS |  |  |  |  |  |  |  |  |
| Wave 1–2: 2014–15 | **3.18 (1.52, 6.65)** | **9.58 (3.43, 26.8)** | 3.10 (0.54, 17.6) | † | 1.15 (0.44, 2.99) | 1.10 (0.37, 3.28) | — | — |
| Wave 2–3: 2015–16 | **2.77 (1.57. 4.91)** | **43.8 (7.48, 256.)** | **14.3 (2.71, 75.2)** | † | 1.86 (0.92, 3.77) | 1.70 (0.56, 5.22) | — | — |
| Wave 3–4: 2016–17 | **3.88 (1.78, 8.45)** | **10.1 (1.60, 64.1)** | 2.23 (0.36, 13.9) | † | 0.97 (0.29, 3.28) | 0.46 (0.14, 1.50) | — | — |
| Wave 4–5: 2017–19 | 2.04 (1.51, 2.77) | **81.2 (17.7, 371.)** | **8.89 (1.67, 47.4)** | † | 0.52 (0.34, 0.79) | 0.69 (0.35, 1.39) | — | — |
| Wave 5–6: 2019–21 | 1.24 (0.87, 1.76) | **34.5 (3.96, 301.)** | 3.05 (0.32, 29.3) | † | 1.15 (0.67, 1.98) | 0.53 (0.20, 1.42) | — | — |
| Wave 6–7: 2021–22 | 1.17 (0.80, 1.72) | **29.3 (6.17, 139.)** | **6.30 (1.18, 33.5)** | † | 0.91 (0.55, 1.50) | 0.90 (0.47, 1.71) | — | — |
| Non-current to cigarette |  |  |  |  |  |  |  |  |
| Wave 1–2: 2014–15 | 1.10 (0.59, 2.04) | **3.13 (1.04, 9.41)** | 1.68 (0.60, 4.69) | **0.24 (0.12, 0.50)** | 1.73 (0.74, 4.03) | 1.69 (0.69, 4.12) | — | — |
| Wave 2–3: 2015–16 | 0.76 (0.47, 1.22) | **4.23 (2.09, 8.58)** | **3.47 (1.72, 7.00)** | **0.44 (0.27, 0.71)** | 0.83 (0.52, 1.33) | 1.38 (0.67, 2.82) | — | — |
| Wave 3–4: 2016–17 | 1.38 (1.00, 1.91) | **2.58 (1.62, 4.12)** | **3.44 (2.45, 4.83)** | **0.37 (0.25, 0.54)** | 0.98 (0.66, 1.46) | 1.09 (0.62, 1.91) | — | — |
| Wave 4–5: 2017–19 | 0.83 (0.57, 1.22) | **2.92 (1.91, 4.45)** | **3.07 (2.04, 4.61)** | **0.41 (0.25, 0.66)** | 0.97 (0.62, 1.52) | **2.22 (1.11, 4.46)** | — | — |
| Wave 5–6: 2019–21 | 0.99 (0.63, 1.54) | 1.19 (0.67, 2.13) | 1.54 (0.92, 2.60) | **0.41 (0.25, 0.66)** | 1.57 (0.94, 2.63) | 1.15 (0.57, 2.32) | — | — |
| Wave 6–7: 2021–22 | 0.92 (0.64, 1.30) | 0.69 (0.34, 1.42) | **1.78 (1.12, 2.83)** | **0.37 (0.24, 0.58)** | **1.61 (1.09, 2.38)** | **1.82 (1.10, 3.02)** | — | — |
| Non-current to ENDS |  |  |  |  |  |  |  |  |
| Wave 1–2: 2014–15 | **2.90 (1.17, 7.20)** | **5.64 (1.48, 21.5)** | 2.39 (0.75, 7.61) | **0.28 (0.08, 0.97)** | 0.53 (0.10, 2.84) | 1.21 (0.28, 5.21) | — | — |
| Wave 2–3: 2015–16 | 1.46 (0.48, 4.41) | **18.1 (2.33, 141)** | 2.22 (0.54, 9.10) | **0.29 (0.12, 0.71)** | 0.90 (0.24, 3.40) | 2.55 (0.75, 8.65) | — | — |
| Wave 3–4: 2016–17 | 1.65 (0.65, 4.17) | **24.1 (7.51, 77.4)** | **4.08 (1.35, 12.4)** | 0.39 (0.08, 1.91) | 0.94 (0.27, 3.24) | 0.50 (0.11, 2.20) | — | — |
| Wave 4–5: 2017–19 | 0.81 (0.48, 1.38) | **13.8 (8.68, 22.0)** | **2.62 (1.54, 4.45)** | **0.32 (0.11, 0.92)** | 0.84 (0.43, 1.64) | 0.76 (0.36, 1.61) | — | — |
| Wave 5–6: 2019–21 | 1.06 (0.61, 1.83) | **18.6 (8.83, 39.2)** | **5.33 (2.78, 10.2)** | **0.20 (0.05, 0.76)** | **0.45 (0.23, 0.90)** | 1.13 (0.40, 3.20) | — | — |
| Wave 6–7: 2021–22 | 0.96 (0.68, 1.34) | **16.3 (9.54, 27.9)** | **4.99 (2.87, 8.69)** | **0.06 (0.02, 0.18)** | 0.88 (0.54, 1.41) | 1.08 (0.53, 2.20) | — | — |
| Cigarette to non-current |  |  |  |  |  |  |  |  |
| Wave 1–2: 2014–15 | 0.95 (0.68, 1.34) | **1.93 (1.14, 3.28)** | 0.94 (0.65, 1.34) | 1.37 (0.90, 2.09) | 1.14 (0.79, 1.64) | 0.78 (0.51, 1.19) | **4.05 (2.83, 5.81)** | — |
| Wave 2–3: 2015–16 | 1.05 (0.81, 1.37) | **1.69 (1.12, 2.55)** | **1.70 (1.16, 2.49)** | 1.17 (0.80, 1.69) | 1.18 (0.86, 1.63) | 0.64 (0.45, 0.93) | **5.47 (4.23, 7.08)** | — |
| Wave 3–4: 2016–17 | 1.15 (0.91, 1.45) | **1.08 (1.14, 2.84)** | 1.06 (0.72, 1.54) | 1.15 (0.80, 1.67) | 1.14 (0.78, 1.67) | 0.78 (0.54, 1.13) | **4.01 (3.06, 5.25)** | — |
| Wave 4–5: 2017–19 | 0.99 (0.78, 1.26) | **1.83 (1.28, 2.61)** | 1.19 (0.88, 1.62) | 0.99 (0.74, 1.33) | 1.05 (0.77, 1.43) | 0.62 (0.44, 0.89) | **4.71 (3.69, 6.01)** | — |
| Wave 5–6: 2019–21 | 1.03 (0.80, 1.33) | 1.49 (0.92, 2.41) | **1.95 (1.37, 2.77)** | 1.35 (0.95, 1.92) | 0.92 (0.60, 1.40) | 0.85 (0.62, 1.16) | **4.72 (3.63, 6.14)** | — |
| Wave 6–7: 2021–22 | 1.01 (0.72, 1.42) | **2.53 (1.39, 4.63)** | **1.53 (1.01, 2.33)** | 0.96 (0.66, 1.41) | 1.32 (0.84, 2.08) | 0.84 (0.57, 1.26) | **6.54 (4.70, 9.09)** | — |
| Cigarette to dual |  |  |  |  |  |  |  |  |
| Wave 1–2: 2014–15 | 1.03 (0.77, 1.37) | **1.71 (1.25, 2.36)** | **1.50 (1.10, 2.06)** | 0.69 (0.46, 1.04) | 0.53 (0.32, 0.88) | 0.26 (0.14, 0.49) | **0.61 (0.43, 0.87)** | — |
| Wave 2–3: 2015–16 | 0.90 (0.67, 1.20) | **2.37 (1.57, 3.58)** | **1.82 (1.21, 2.74)** | 0.65 (0.41, 1.02) | 0.54 (0.33, 0.88) | 0.41 (0.24, 0.70) | **0.64 (0.44, 0.94)** | — |
| Wave 3–4: 2016–17 | 1.01 (0.76, 1.35) | **2.10 (1.43, 3.07)** | **1.66 (1.24, 2.22)** | **0.36 (0.24, 0.55)** | 0.85 (0.55, 1.31) | 0.53 (0.30, 0.92) | **0.62 (0.42, 0.91)** | — |
| Wave 4–5: 2017–19 | 1.08 (0.87, 1.35) | **3.30 (2.48, 4.40)** | **1.80 (1.27, 2.55)** | **0.44 (0.28, 0.69)** | 0.52 (0.35, 0.78) | 0.45 (0.29, 0.68) | 0.82 (0.61, 1.12) | — |
| Wave 5–6: 2019–21 | 0.61 (0.47, 0.78) | **4.53 (3.26, 6.31)** | **1.97 (1.45, 2.67)** | **0.26 (0.16, 0.41)** | 0.78 (0.50, 1.21) | 0.61 (0.36, 1.04) | 1.26 (0.87, 1.84) | — |
| Wave 6–7: 2021–22 | 0.86 (0.64, 1.14) | **3.38 (2.21, 5.17)** | **1.75 (1.30, 2.36)** | **0.27 (0.18, 0.42)** | 0.72 (0.46, 1.12) | 0.66 (0.41, 1.07) | 1.43 (0.98, 2.11) | — |
| ENDS to non-current |  |  |  |  |  |  |  |  |
| Wave 1–2: 2014–15 | 1.62 (0.85, 3.09) | 0.74 (0.26, 2.12) | 1.04 (0.44, 2.45) | 0.85 (0.34, 2.11) | 1.70 (0.73, 3.98) | **2.90 (1.14, 7.40)** | — | **4.33 (2.44, 7.68)** |
| Wave 2–3: 2015–16 | 0.84 (0.51, 1.40) | 1.58 (0.70, 3.57) | 1.08 (0.47, 2.46) | **0.34 (0.13, 0.89)** | **2.07 (1.04, 4.09)** | **3.39 (1.76, 6.52)** | — | **2.90 (1.47, 5.72)** |
| Wave 3–4: 2016–17 | 0.63 (0.41, 0.96) | 1.16 (0.62, 2.16) | 0.76 (0.44, 1.30) | 0.47 (0.20, 1.11) | 1.59 (0.89, 2.83) | 1.45 (0.79, 2.65) | — | **5.35 (3.35, 8.56)** |
| Wave 4–5: 2017–19 | 0.94 (0.65, 1.37) | 1.12 (0.66, 1.90) | 1.22 (0.70, 2.13) | 1.19 (0.64, 2.21) | 1.22 (0.74, 2.03) | 1.27 (0.69, 2.35) | — | **4.28 (2.77, 6.61)** |
| Wave 5–6: 2019–21 | 1.19 (0.89, 1.60) | 1.30 (0.84, 1.99) | **1.74 (1.13, 2.68)** | 0.93 (0.46, 1.90) | 0.79 (0.50, 1.26) | 1.68 (0.98, 2.88) | — | **2.71 (2.03, 3.60)** |
| Wave 6–7: 2021–22 | 0.85 (0.64, 1.12) | 1.17 (0.77, 1.79) | 1.16 (0.69, 1.96) | 0.48 (0.21, 1.11) | 1.40 (0.96, 2.05) | 1.51 (0.86, 2.65) | — | **2.62 (1.99, 3.45)** |
| ENDS to dual |  |  |  |  |  |  |  |  |
| Wave 1–2: 2014–15 | 1.05 (0.56, 1.99) | 1.62 (0.57, 4.60) | 1.03 (0.36, 2.94) | 1.17 (0.43, 3.17) | 0.13 (0.01, 2.02) | 0.58 (0.07, 4.62) | — | 1.16 (0.52, 2.56) |
| Wave 2–3: 2015–16 | 1.30 (0.67, 2.51) | 1.00 (0.38, 2.62) | 1.29 (0.61, 2.73) | **0.32 (0.11, 0.94)** | 0.77 (0.31, 1.95) | 0.31 (0.03, 3.65) | — | 0.87 (0.34, 2.23) |
| Wave 3–4: 2016–17 | 1.18 (0.68, 2.07) | 1.04 (0.53, 2.02) | 0.45 (0.19, 1.05) | 0.63 (0.29, 1.40) | 1.53 (0.74, 3.16) | 0.55 (0.14, 2.11) | — | 0.88 (0.49, 1.57) |
| Wave 4–5: 2017–19 | 1.02 (0.60, 1.72) | 0.84 (0.40, 1.75) | 1.58 (0.73, 3.44) | 0.48 (0.15, 1.53) | 1.22 (0.61, 2.45) | 0.21 (0.01, 6.50) | — | 0.53 (0.20, 1.41) |
| Wave 5–6: 2019–21 | 1.22 (0.62, 2.39) | 1.28 (0.54, 3.06) | 1.10 (0.43, 2.80) | 0.33 (0.07, 1.72) | 0.42 (0.06, 2.70) | 2.22 (0.55, 9.02) | — | 0.56 (0.26, 1.23) |
| Wave 6–7: 2021–22 | 1.34 (0.65, 2.76) | 0.64 (0.25, 1.62) | 1.08 (0.41, 2.86) | 0.66 (0.18, 2.36) | 0.52 (0.23, 1.17) | 1.51 (0.43, 5.31) | — | 1.55 (0.75, 3.18) |
| Dual to cigarette |  |  |  |  |  |  |  |  |
| Wave 1–2: 2014–15 | 0.80 (0.55, 1.16) | 1.28 (0.82, 1.97) | 1.35 (0.91, 1.99) | 1.30 (0.78, 2.18) | 1.53 (0.87, 2.69) | 1.08 (0.61, 1.91) | 0.72 (0.43, 1.19) | **1.53 (1.09, 2.13)** |
| Wave 2–3: 2015–16 | 0.97 (0.72, 1.31) | 1.16 (0.77, 1.76) | 0.98 (0.67, 1.42) | 0.81 (0.55, 1.20) | 1.31 (0.79, 2.16) | 0.64 (0.33, 1.24) | **0.64 (0.47, 0.87)** | **1.98 (1.45, 2.71)** |
| Wave 3–4: 2016–17 | 1.22 (0.95, 1.57) | 1.42 (0.99, 2.05) | 0.94 (0.67, 1.33) | 0.91 (0.58, 1.42) | 0.89 (0.51, 1.55) | 1.26 (0.67, 2.38) | 0.73 (0.51, 1.04) | **1.77 (1.30, 2.41)** |
| Wave 4–5: 2017–19 | 0.89 (0.65, 1.21) | 0.76 (0.49, 1.17) | 0.92 (0.61, 1.39) | 1.39 (0.91, 2.14) | 1.09 (0.67, 1.79) | 0.93 (0.54, 1.61) | **0.62 (0.40, 0.96)** | **1.53 (1.10, 2.13)** |
| Wave 5–6: 2019–21 | 1.13 (0.84, 1.52) | 0.64 (0.43, 0.97) | 0.66 (0.47, 0.94) | **0.55 (0.34, 0.90**) | 0.93 (0.53, 1.64) | 1.14 (0.65, 2.00) | 0.76 (0.52, 1.10) | **1.84 (1.32, 2.57)** |
| Wave 6–7: 2021–22 | 1.35 (0.87, 2.09) | 0.66 (0.35, 1.23) | 0.74 (0.46, 1.19) | 0.83 (0.45, 1.51) | 0.83 (0.39, 1.78) | 1.22 (0.57, 2.59) | 0.60 (0.32, 1.12) | 1.38 (0.87, 2.18) |
| Dual to ENDS |  |  |  |  |  |  |  |  |
| Wave 1–2: 2014–15 | 0.96 (0.42, 2.19) | 1.42 (0.53, 3.82) | 1.60 (0.67, 3.83) | 1.72 (0.43, 6.92) | 1.42 (0.42, 4.80) | 0.74 (0.07, 7.45) | 1.92 (0.91, 4.04) | 0.51 (0.22, 1.17) |
| Wave 2–3: 2015–16 | 1.05 (0.60, 1.85) | 1.28 (0.65, 2.53) | 0.89 (0.43, 1.85) | **0.34 (0.12, 0.95)** | 0.72 (0.17, 3.07) | 1.54 (0.52, 4.52) | 1.64 (0.94, 2.87) | **0.31 (0.15, 0.66)** |
| Wave 3–4: 2016–17 | 0.96 (0.57, 1.62) | 1.15 (0.50, 2.65) | 0.96 (0.47, 1.98) | 0.96 (0.30, 3.06) | 1.31 (0.56, 3.05) | 0.45 (0.07, 2.75) | **4.54 (2.41, 8.53)** | 1.31 (0.69, 2.48) |
| Wave 4–5: 2017–19 | 1.11 (0.68, 1.79) | **4.54 (2.41, 8.55)** | 3.26 (1.58, 6.74) | 1.85 (0.72, 4.78) | 0.67 (0.24, 1.82) | 1.35 (0.62, 2.94) | **2.00 (1.32, 3.03)** | 0.60 (0.35, 1.02) |
| Wave 5–6: 2019–21 | 0.75 (0.48, 1.17) | **2.64 (1.59, 4.38)** | 1.37 (0.84, 2.24) | 0.77 (0.36, 1.65) | 1.23 (0.71, 2.12) | 1.23 (0.56, 2.69) | **1.70 (1.08, 2.67)** | **0.39 (0.26, 0.59)** |
| Wave 6–7: 2021–22 | 0.94 (0.64, 1.37) | 1.43 (0.82, 2.50) | 1.69 (0.95, 2.99) | 0.54 (0.17, 1.71) | 1.06 (0.59, 1.88) | 1.12 (0.63, 2.00) | **3.07 (2.06, 4.57)** | **0.59 (0.35, 0.98)** |
| †Transition in the numerator is negligible | | | | | | | | |
